# Tau spreading is driven by neuronal connectivity in primary tauopathies - evidence from tau-PET and histopathology

**DOI:** 10.1101/2021.08.16.21261523

**Authors:** Nicolai Franzmeier, Matthias Brendel, Leonie Beyer, Gabor Kovacs, Thomas Arzberger, Carolin Kurz, Gesine Respondek, Milica Jecmenica Lukic, Davina Biel, Anna Rubinski, Lukas Frontzkowski, Anika Finze, Carla Palleis, Emanuel Joseph, Endy Weidinger, Sabrina Katzdobler, Mengmeng Song, Gloria Biechele, Maike Kern, Maximilian Scheifele, Boris-Stephan Rauchmann, Robert Perneczky, Michael Rullman, Marianne Patt, Andreas Schildan, Henryk Barthel, Osama Sabri, Jost J. Rumpf, Matthias L. Schroeter, Joseph Classen, Victor Villemagne, John Seybl, Andrew W. Stephens, Edward B. Lee, David G. Coughlin, Armin Giese, Murray Grossman, Corey T. McMillan, Ellen Gelpi, Laura Molina-Porcel, Yaroslau Compta, John C. van Swieten, Laura Donker Laat, Claire Troakes, Safa Al-Sarraj, John L. Robinson, Sharon X. Xie, David J. Irwin, Sigrun Roeber, Jochen Herms, Mikael Simons, Peter Bartenstein, Virginia M. Lee, John Q. Trojanowski, Johannes Levin, Günter U. Höglinger, Michael Ewers, For the German Imaging Initiative for Tauopathies (GII4T) & the Alzheimer’s Disease Neuroimaging Initiative

**Affiliations:** Institute for Stroke and Dementia Research, Klinikum der Universität München, Ludwig-Maximilians-Universität LMU, Munich, Germany; Department of Nuclear Medicine, University Hospital of Munich, LMU Munich, Munich, Germany; Munich Cluster for Systems Neurology (SyNergy), Munich, Germany; Center for Neurodegenerative Disease Research (CNDR), Institute On Aging and Department of Pathology & Laboratory Medicine, University of Pennsylvania, Philadelphia, USA; Tanz Centre for Research in Neurodegenerative Disease (CRND) and Department of Laboratory Medicine and Pathobiology, University of Toronto, Toronto, Canada; Laboratory Medicine Program and Krembil Brain Institute, University Health Network, Toronto, ON, Canada; German Center for Neurodegenerative Diseases (DZNE), Munich, Germany; Department of Psychiatry and Psychotherapy, University Hospital, LMU Munich, Munich, Germany; Center for Neuropathology and Prion Research, LMU Munich, Munich, Germany; Department of Neurology, Hannover Medical School, Hannover, Germany; Clinic of Neurology, University Clinical Center of Serbia, Belgrade, Republic of Serbia; Department of Neurology, University Hospital of Munich, LMU Munich, Munich, Germany; Department of Radiology, University Hospital of Munich, LMU Munich, Munich, Germany; Ageing Epidemiology Research Unit (AGE), School of Public Health, Imperial College, London, United Kingdom; Department of Nuclear Medicine, University Hospital Leipzig, Leipzig, Germany; Department of Neurology, University Hospital Leipzig, Leipzig, Germany; Clinic for Cognitive Neurology, University of Leipzig, Leipzig, Germany, Max- Planck-Institute of Human Cognitive and Brain Sciences, Leipzig, Germany; Department of Molecular Imaging & Therapy, Austin Health, Heidelberg, VIC, Australia; Department of Psychiatry, University of Pittsburgh, Pittsburgh, PA, USA; Department of Medicine, Austin Health, The University of Melbourne, Melbourne, VIC, Australia; InviCRO, LLC, Boston, MA, United States of America; Molecular Neuroimaging, A Division of inviCRO, New Haven, CT, United States of America; Life Molecular Imaging GmbH, Berlin, Germany; Frontotemporal Degeneration Center, University of Pennsylvania, Philadelphia, USA; Department of Neurosciences, University of California, La Jolla, San Diego, CA, USA; Department of Neurology, University of Pennsylvania, Philadelphia, USA; Neurological Tissue Bank and Neurology Department, Hospital Clínic de Barcelona, Universitat de Barcelona, IDIBAPS, CERCA, Barcelona, Catalonia, Spain; Alzheimer’s disease and other cognitive disorders unit. Neurology Service, Hospital Clínic, Institut d’Investigacions Biomediques August Pi i Sunyer (IDIBAPS), University of Barcelona, Barcelona, Spain; Parkinson’s Disease & Movement Disorders Unit, Hospital Clínic / IDIBAPS / CIBERNED (CB06/05/0018-ISCIII), / European Reference Network for Rare Neurological Diseases (ERN-RND) / Institut de Neurociències (Maria de Maeztu Center), Universitat de Barcelona, Barcelona, Catalonia, Spain; Department of Neurology, Erasmus Medical Centre, Rotterdam, The Netherlands; Department Clinical Genetics, Erasmus Medical Center, Rotterdam, The Netherlands; London Neurodegenerative Diseases Brain Bank, Institute of Psychiatry, Psychology and Neuroscience, Kings College London, London, UK; Department of Biostatistics, Epidemiology and Informatics, University of Pennsylvania, Philadelphia, USA; Department of Neurology, Technical University of Munich, Munich, Germany

## Abstract

Tau pathology is the main driver of neuronal dysfunction in 4-repeat tauopathies (4RT), including cortico-basal degeneration and progressive supranuclear palsy (PSP). Tau is assumed to spread prion-like across connected neurons, but the mechanisms of tau propagation are largely elusive in 4RTs, characterized not only by neuronal but also by astroglial and oligodendroglial tau accumulation. Here, we assessed whether connectivity drives 4R-tau spreading patterns by combining resting-state fMRI connectomics with both 2^nd^ generation ^18^F- PI-2620 tau-PET in 46 patients with clinically diagnosed 4RTs and post-mortem cell-type-specific regional tau assessments from two independent PSP samples (n=97/96). We found that inter-regional connectivity was associated with higher inter-regional correlation of both tau-PET and post-mortem tau levels in 4RTs. In regional cell-type specific post-mortem tau assessments, this association was stronger for neuronal than for astroglial or oligodendroglial tau, suggesting that connectivity is primarily associated with trans-neuronal tau spread. Using tau-PET we found that patient-level tau patterns can be predicted by the connectivity of subcortical tau epicenters. Together, the current study provides combined in vivo tau-PET and histopathological evidence for brain connectivity as a key mediator of trans-neuronal tau spreading in 4RTs.

## INTRODUCTION

Progressive supranuclear palsy (PSP) and cortico-basal degeneration (CBD) are primary 4- repeat (4R) tauopathies characterized by glial and neuronal 4R-tau inclusions, manifesting as progressive movement and cognitive disorders.^1–4^ In PSP and CBD, 4R-tau pathology accumulates initially in the brainstem and subcortex, with subsequent cortical manifestation at more advanced disease stages.^5, 6^ In fact, PSP and CBD share core genetic,^7, 8^ biochemical and neuropathologic features,^3, 9^ hence there is debate as to whether or not they should be regarded as a single pathophysiological disease entity.^10, 11^

Post-mortem studies have reported a strong clinico-pathological correlation between 4R-tau deposition patterns and clinical phenotype: PSP most commonly presents as Richardson’s syndrome (PSP-RS, i.e. a combination of postural instability and ocular motor deficits), associated with brainstem and subcortical tau followed by progressive tau accumulation in parietal and motor cortices at later disease stages.^12, 13^ Further variant predominance types include, among others, PSP with speech/language disorder with left inferior frontal tau aggregation or PSP with cognitive/behavioral presentation with fronto-temporal tau pathology, suggesting that clinical variability is driven by spatially heterogeneous tau patterns.^13–15^ In accord with this concept, CBD patients often present with mixed cortical/movement disorders termed cortico-basal syndrome (CBS), associated with tau accumulation in the motor cortex, brainstem, subthalamic nucleus, and striatum, yet clinical variants present as progressive aphasia, frontal-behavioral syndrome or Richardson’s syndrome.^16, 17^ Since 4R tau deposition patterns in PSP and CBD are considered key determinants of disease phenotype and progression, a detailed understanding of the mechanisms that facilitate tau spreading is of pivotal clinical interest.

There is converging preclinical evidence suggesting cell-to-cell tau pathology transmission across functionally interconnected neurons:^18^ First, pathological tau strains obtained from patients with primary or secondary tauopathies have been shown to induce template-based misfolding of physiological tau, suggesting “prion-like” tau propagation.^19, 20^ Second, injection of pathological tau seeds in mouse brains triggers tau spread to regions anatomically connected to the injection site rather than tau diffusion to spatially adjacent regions.^20–24^ Third, synapses and neuronal activity are considered to be key drivers of tau spreading,^25, 26^ where opto- genetically enhanced activity of tau harboring neurons is associated with accelerated trans-synaptic tau spreading in vitro and in vivo.^27^ Collectively, these pre-clinical findings provide strong support for neuronal connectivity as a key route for tau spreading in tauopathies.

However, it remains unclear whether tau pathology progresses preferentially between connected brain regions in human 4R tauopathies. This spreading mechanism needs to be specifically questioned in 4R tauopathies since tau is not exclusively present in neurons, but also in astroglia and oligodendroglia.^13^ To this end, we combined in vivo tau-PET and post-mortem tau assessments from 4R tauopathy patients with fMRI-based connectivity assessments obtained in a healthy reference sample. Using ^18^F-PI-2620 PET for tau imaging in clinically diagnosed CBS (n=24) and PSP-RS patients (n=22), we tested if i) functionally connected brain regions show correlated ^18^F-PI-2620 PET levels and whether ii) brain-wide tau-PET uptake patterns can be predicted by the functional connectivity pattern of subcortical epicenters with highest tau pathology. In CBS, which is typically associated with more widespread cortical tau,^28^ we further tested whether subthreshold cortical Aβ levels may enhance the spread of tau-PET from subcortical tau epicenters to the cortex. This analysis was motivated by previous reports showing clinical and pathophysiological overlap between CBS and AD,^29^ where a substantial number of clinical CBS cases had co-occurring Aβ pathology,^28, 30^ i.e. a key driver of cortical tau spreading in AD.^31^ In a key validation step, we translated the in vivo tau-PET analyses to two independent post-mortem samples (Munich-European consortium/collection): n=97, University of Pennsylvania [UPENN]: n=96) with confirmed PSP pathology and gold-standard regional histopathological 4R tau assessments. Inclusion of this post-mortem sample allowed cell-type specific stratification of tau pathology (i.e. neuronal, astroglial and oligodendroglial tau) to specifically test the hypothesis that inter-regional connectivity is associated with the spreading of neuronal tau pathology. By combining cell-type-specific post-mortem tau assessments with atlas-based functional connectivity data, we tested the hypothesis that higher functional connectivity between post-mortem sampled brain regions is associated with more correlated tau levels, and that this association is primarily driven by neuronal trans-synaptic tau spreading.

## RESULTS

We obtained tau-PET in 46 patients with clinically suspected 4R tauopathies, including 24 CBS patients, 22 PSP-RS patients and 15 cognitively normal controls (CN). All subjects underwent 3T structural MRI and ^18^F-PI-2620 PET imaging.^32^ Aβ-positive CBS patients (as determined by cerebrospinal fluid Aβ levels or amyloid-PET, which was available for all CBS cases) were considered to have underlying 3R/4R tau AD pathology and were therefore excluded from the current study.^33^ Sample characteristics are presented in table 1. Group-average ^18^F-PI-2620 SUVR maps intensity normalized to the inferior cerebellar grey matter are shown in Fig.1 for CN (Fig.1A), PSP-RS (Fig.1B) and CBS (Fig.1C). For neuropathological validation, we included post-mortem tau assessments from two independent samples (Munich-European consortium/collection sample, n=97; UPENN sample, n=96) with histopathologically confirmed PSP, defined as the presence of neurofibrillary tau tangles (NFT) in the subthalamic nucleus, substantia nigra and globus pallidus together with tufted astrocytes in the striatum and frontal cortex.^3, 13^ For functional connectivity, we included resting-state fMRI data of 69 cognitively normal subjects from the Alzheimer’s disease neuroimaging initiative (ADNI) cohort without evidence of amyloid or tau pathology as indicated by ^18^F-florbetapir amyloid-PET and ^18^F-flortaucipir tau-PET.^34, 35^

**Figure 1:**
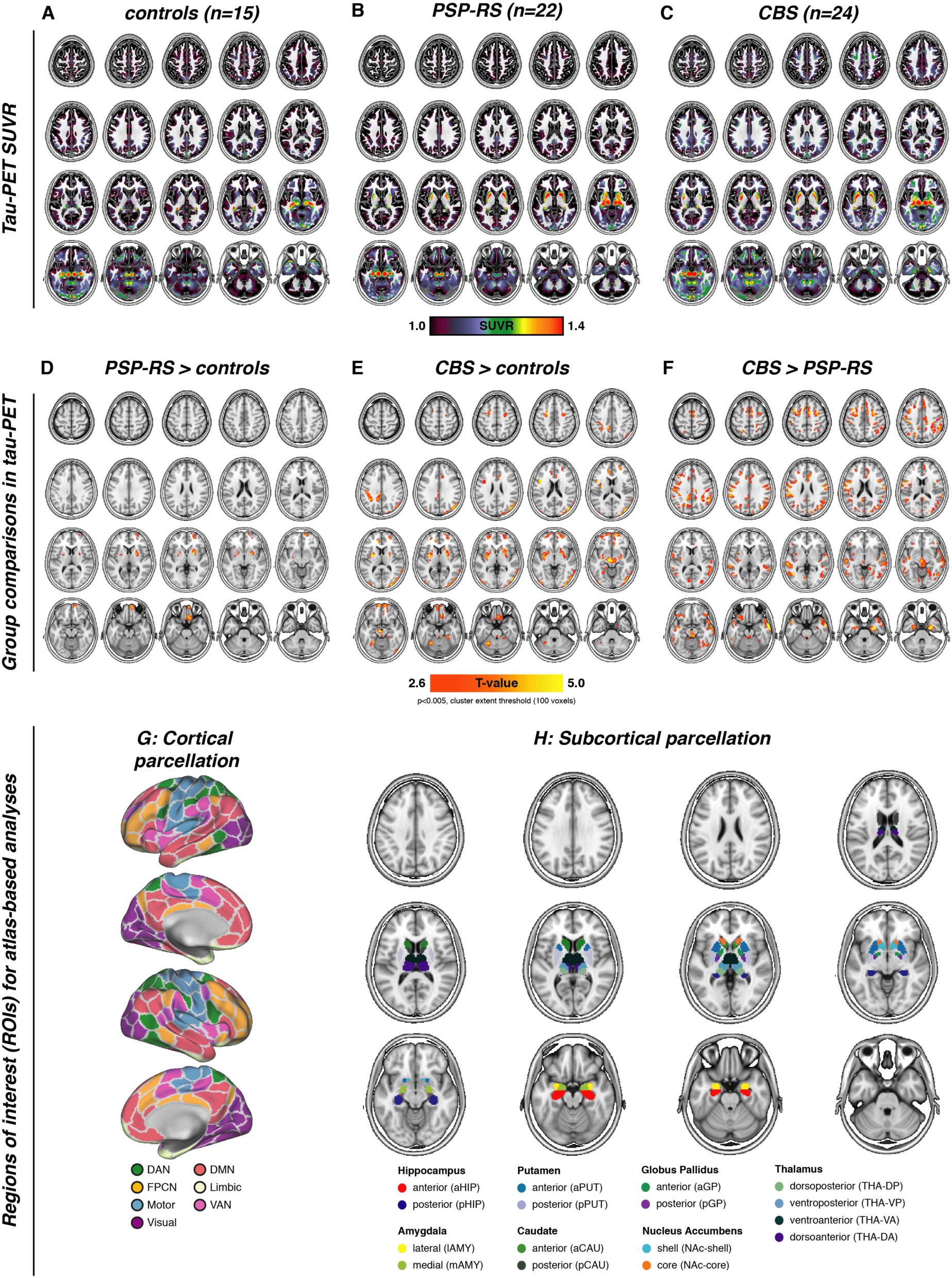
Group-average maps of tau-PET SUVRs (i.e. intensity normalized to the inferior cerebellar grey) for controls (A), PSP-RS (B) and CBS patients (C). Voxel-wise group comparisons were conducted to compare tau- PET SUVRs between PSP-RS vs. controls (D), CBS vs. controls (E) and CBS vs. PSP-RS, at a voxel threshold of p<0.005 with a cluster size of at least 100 spatially contiguous voxels. Illustration of the 200 ROI cortical (G) and 32 ROI subcortical (H) brain atlases that were used for all tau-PET vs. connectivity analyses.

**Table 1:** Tau-PET sample

|  | **Controls (n=15)** | **PSP-RS (n=22)** | **CBS (n=24)** | **P** |
| --- | --- | --- | --- | --- |
| Age | 62.67±8.97<sup>b</sup> | 72.07±6.4<sup>a</sup> | 67.71±8.46 | 0.001 |
| Sex (f/m) | 9/6 | 10/18 | 14/10 | 0.175 |
| Disease Duration<br>(months) | NA | 39.48±25.01 | 28.86±21.48 | 0.143 |

a=significantly different from controls (p<0.05), b=significantly different from PSP-RS (p<0.05), c=significantly different from CBS (p<0.05)

### Elevated tau-PET binding in PSP-RS/CBS

First, we assessed ^18^F-PI-2620 PET binding in CBS and PSP-RS patients vs. CN, using voxel-wise ANCOVAs, controlling for age, sex and study site (alpha-threshold=0.005, cluster-extent threshold>100 voxels). Compared to CN, PSP-RS patients showed higher subcortical and inferior frontal tau-PET binding (Fig.1D), congruent with previous work in a partly overlapping sample.^32^ In CBS,^36^ we also found significantly elevated subcortical, brainstem and inferior frontal^18^F-PI-2620 PET binding but also more widespread occipital, frontal and parietal elevations of ^18^F-PI-2620 PET binding (Fig.1E). When comparing ^18^F-PI-2620 PET between CBS and PSP (Fig.1F), we found higher cortical ^18^F-PI-2620 PET binding in CBS in the midbrain, entorhinal and lateral temporal cortex, motor cortex, anterior cingulate, frontal eye fields and parietal cortex. In line with histopathological studies,^3^ these analyses suggest elevated tau in patients with suspected 4R tauopathies vs. CN, particularly in the subcortex, with more widespread cortical tau in CBS vs. PSP-RS.^28^

### Functional connectivity is associated with correlated tau-PET binding in CBS/PSP-RS

Next, we tested the major hypothesis whether inter-regional connectivity is associated with inter-regionally correlated tau accumulation in CBS or PSP-RS. To this end, we parcellated patient-specific ^18^F-PI-2620 PET images into 232 regions-of-interests (ROIs) by combining non-overlapping functional MRI-based parcellations of the cortex (200 ROIs, Fig.1G)^37^ and subcortex (32 ROIs, Fig.1H).^38^ Primary analyses focused on the subcortex (32 ROIs, Fig.1G), which showed consistently elevated ^18^F-PI-2620 PET binding in CBS and PSP-RS (see Fig.1E&F). For secondary analyses we also included the neocortex (i.e. 32 subcortical plus 200 cortical ROIs, Fig.1G), which was less consistently affected by elevated ^18^F-PI-2620 PET binding in PSP-RS/CBS (see Fig.1E&F).

We determined the covariance in inter-regional ^18^F-PI-2620 PET binding within CBS and PSP-RS groups, defined as partial correlation of ROI-based ^18^F-PI-2620 PET SUVRs, accounting for age, sex and imaging site as confounds (methods illustrated in Fig.2A, for subcortical tau covariance matrices see Fig.2B for PSP-RS and Fig.2C for CBS). For functional connectivity, we applied the same 232 ROI-based parcellation to pre-processed resting-state fMRI data of 69 cognitively normal ADNI subjects. Based on this healthy control sample, we determined a group-average functional connectivity template (see Fig.2D). Using linear regression, we next assessed whether higher inter-regional functional connectivity was associated with higher inter-regional covariance of ^18^F-PI-2620 PET binding, controlling for between-ROI Euclidean distance to ensure that associations were not driven by spatial proximity. For our primary analyses on the subcortex (i.e. where tau-PET was significantly elevated in PSP-RS and CBS compared to controls), we found the expected associations between higher functional connectivity and higher ^18^F-PI-2620 PET covariance in PSP-RS (β=0.616, p<0.001, Fig.3A) and CBS (β=0.561, p<0.001, Fig.3B). To test the robustness of these findings, we recomputed these models 1000 times generating for each trial a new connectivity null-model (i.e. shuffled connectivity matrix with preserved weight- and degree distribution), yielding a distribution of null-model derived β-values (Figs. 3A&B, beeswarm panels). Comparing the actual β-values estimated using the observed “true” connectivity matrix with the null-distributions using an exact test (i.e. determine the probability of null-distribution derived β-values surpassing the true β-value), yielded p-values <0.001 for CBS and PSP-RS. Together, these analyses support the view that functionally connected subcortical regions show covarying tau-PET levels in CBS/PSP-RS. We obtained consistent results when extending these analyses to the whole brain (Figs.1G&H), where higher connectivity was again associated with higher ^18^F-PI-2620 PET covariance in PSP-RS (Fig.3C, β=0.450, p<0.001) and CBS (Fig.3D, β=0.402, p<0.001), controlling for inter-regional Euclidean distance. Exact tests using shuffled connectivity data yielded consistent results (p<0.001, Figs.3C&D, beeswarm panels). We next tested whether this association was consistent across subcortico-cortical and cortico-cortical connections. Again, associations between connectivity and ^18^F-PI-2620 PET covariance were found for cortico-cortical (PSP-RS: β=0.397, p<0.001; CBS: β=0.397, p<0.001) and subcortico-cortical connections (PSP-RS: β=0.329, p<0.001; CBS: β=0.379, p<0.001). Together, these findings of correlated tau-PET among functionally connected regions support the idea that connectivity shapes tau spreading patterns in 4R tauopathies.

**Figure 2:**
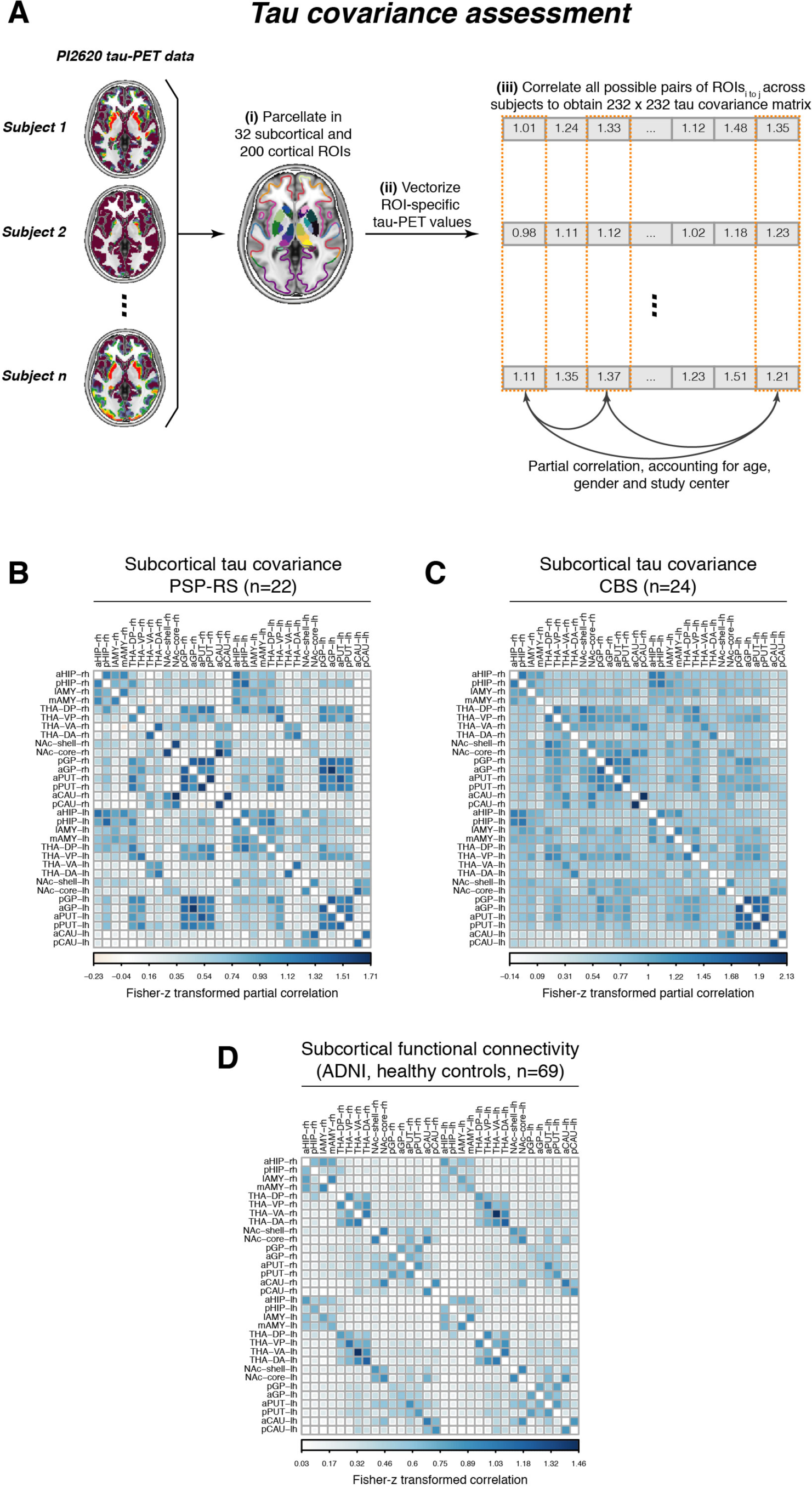
Flow-chart illustrating the assessment of tau covariance (A). Subject-level tau-PET data was parcellated into 200 cortical and 32 subcortical ROIs (i), mean tau-PET was extracted for each ROI, vectorized to 232 element vectors and concatenated across subjects (ii). Fisher-z transformed partial-correlations between inter-regional tau-PET SUVRs were determined for each group (i.e. PSP-RS and CBS), accounting for age, sex and study site (iii). The resulting tau covariance matrices for the subcortical brain parcellation which was used for primary analyses is shown for PSP-RS (B) and CBS (C) patients. For the same ROIs, group-average functional connectivity was computed based on resting-state fMRI of 69 cognitively normal, amyloid and tau negative ADNI participants.

**Figure 3:**
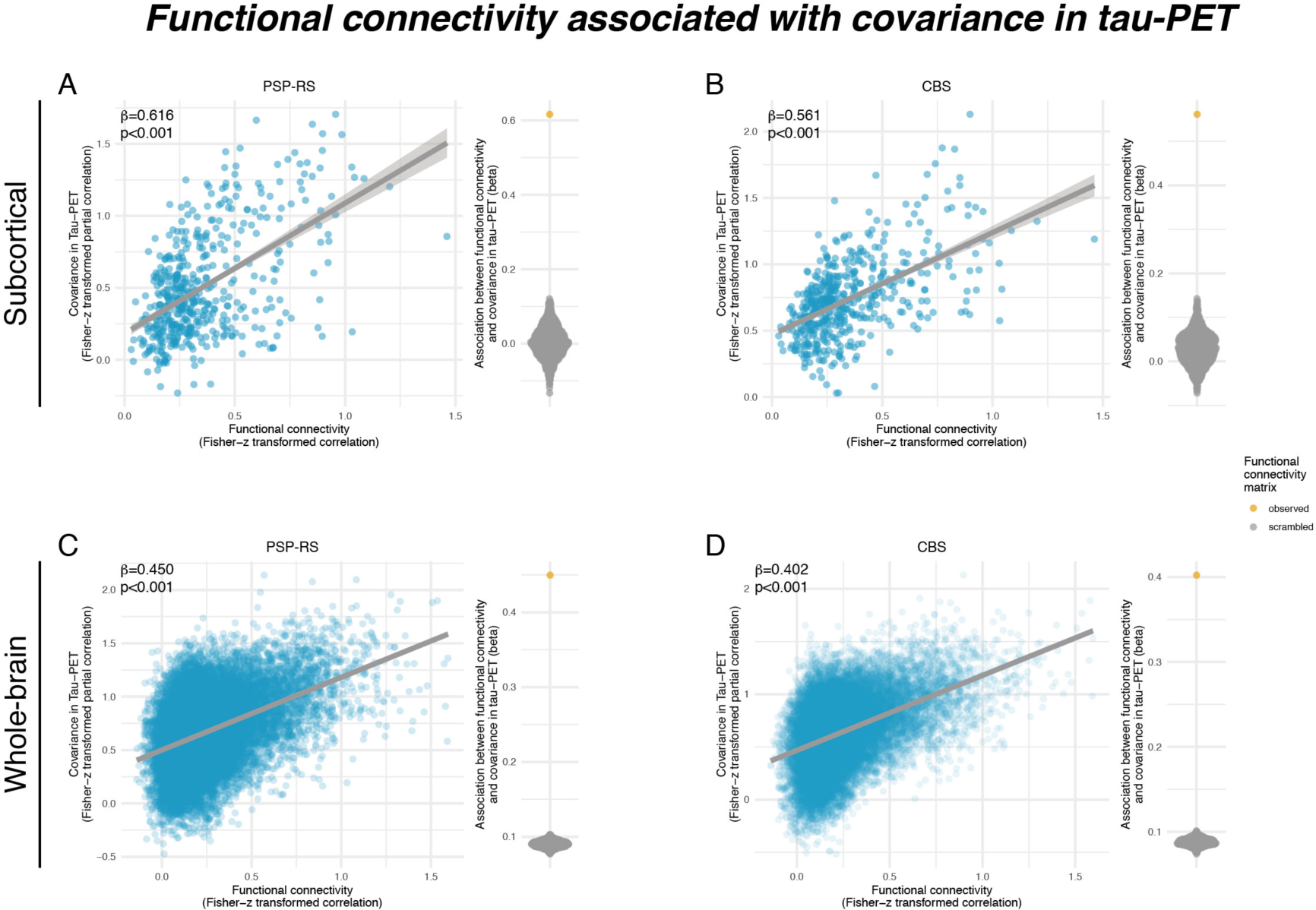
Scatterplots illustrating the association between functional connectivity and covariance in 18-F- PI2620-PET among subcortical regions in PSP-RS (A) and CBS groups (B), as well as among subcortical and cortical regions for PSP-RS (C) and CBS groups (D). Standardized beta- and p-values were derived from linear regression controlling for Euclidean distance between ROIs. Beeswarm plots illustrate the distribution of standardized beta-values derived from repeating the analysis 1000 times using scrambled connectomes with preserved weight- and degree-distribution (grey points) vs. the beta value derived from the association with the actual observed connectivity matrix that is illustrated in the scatterplot (yellow point).

### Functional connectivity predicts tau-PET binding in PSP-RS/CBS

We next determined whether group-average ^18^F-PI-2620 PET patterns follow the connectivity pattern of subcortical tau epicenters (i.e. 20% of brain regions with highest tau-PET). We found that for epicenter regions with highest ^18^F-PI-2620 PET binding, higher seed-based functional connectivity was associated with higher tau-PET binding in strongly connected subcortical regions in both PSP-RS (Fig.4A, β=0.880, p<0.001) and CBS (Fig.4B, β=0.933, p<0.001), controlling for between-ROI Euclidean Distance. In turn, for coldspot regions with lowest ^18^F- PI-2620 PET binding, higher seed-based functional connectivity was associated with lower subcortical ^18^F-PI-2620 PET binding in strongly connected regions in PSP-RS (Fig.4C, β=- 0.613, p<0.001) and CBS (Fig.4D, β=-0.617, p<0.001), controlling for between-ROI Euclidean distance. This result pattern was reflected in a strong positive correlation between the seed ROIs ^18^F-PI-2620 PET binding and their functional connectivities’ predictive weight (i.e. regression derived β-value) on ^18^F-PI-2620 PET binding in remaining ROIs (PSP-RS: β=0.929, p<0.001, Fig.4E; CBS: β=0.937, p<0.001, Fig.4F). A fully congruent result pattern was detected when extending this approach to the whole brain (see Figs. 4G-L). Together, these findings suggest that regions with high tau-PET levels are primarily interconnected with regions also harboring high tau, whereas regions with low tau are primarily connected to other low tau regions.

**Figure 4:**
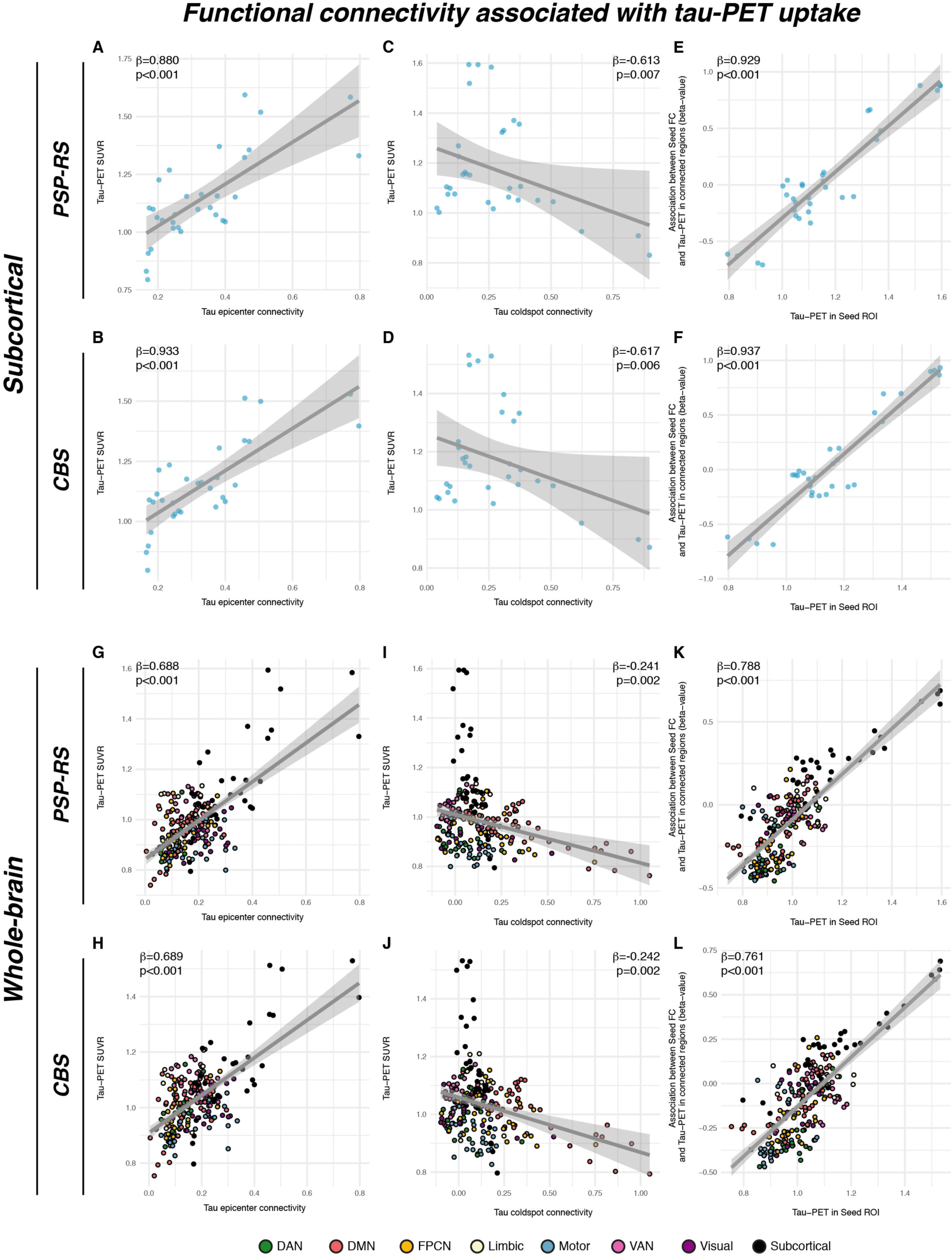
Associations between group-average subcortical 18-F-Pi2620-PET data and seed-based functional connectivity of tau epicenters (i.e. regions with highest group-average tau) in PSP-RS (A) and CBS (B), illustrating that regions with high connectivity to the tau epicenter show high tau-PET. The same association was plotted for tau coldspots (i.e. regions with lowest tau-PET) for PSP-RS (C) and CBS (D), illustrating that regions closely connected to the tau coldspots show also low tau-PET. Standardized beta- and p-values were derived from linear regression controlling for Euclidean distance between ROIs. The analysis was repeated for all ROIs, and the respective seed ROIs tau-PET uptake was plotted against the regression derived beta-value, showing that seed regions with higher tau-PET show a positive association between seed-based connectivity and tau-PET in connected regions, whereas regions with lower tau-PET show a negative association between seed-based connectivity and tau-PET in connected regions in PSP (E) and CBS (F). These findings indicate that seed ROIs are preferentially connected to other regions with similar tau-PET levels. All analyses were repeated including using the combined set of 200 cortical and 32 subcortical ROIs, showing a fully consistent result pattern across the entire brain (G-L).

### Connectivity of patient-specific tau epicenters predicts individual tau-PET binding in 4R tauopathy patients

As shown by the group-level analyses, epicenter regions with highest tau-PET binding were most strongly connected to other regions with high ^18^F-PI-2620 PET in PSP-RS (Fig. 4A&G) and CBS (Fig. 4B&H). Next, we extended this analysis to the subject level, i.e. we defined each PSP-RS and CBS patients’ tau epicenter as those ∼20% of subcortical ROIs with the highest ^18^F-PI-2620 PET SUVR.^39^ Adopting our previously established approach,^39^ the remaining non-epicenter subcortical ROIs were grouped for each subject into 4 quartiles depending on the mean connectivity strength to a given subjects’ tau epicenter (Q1=strongest connectivity to the tau epicenter vs. Q4=weakest connectivity to the tau epicenter) as illustrated in Fig.5A. We expected a gradient of ^18^F-PI-2620 PET decrease from tau epicenters across functionally connected regions (i.e. highest tau in Q1, vs. lowest tau in Q4). Using linear mixed models, we could confirm that connectivity strength to the epicenter was predictive of ^18^F-PI-2620 PET binding in connected Q1-Q4 regions for PSP-RS (b-value/standard error [b/SE]=0.097/0.015, p<0.001, Fig.5B) and CBS (Fig.5D, b/SE=0.086/0.013, p<0.001). As hypothesized, highest ^18^F- PI-2620 PET binding was found in Q1, which is most strongly connected to the tau epicenter, whereas tau-PET levels gradually decreased to Q4, which is only weakly connected to the tau epicenter. Linear mixed models were controlled for age, sex, study site, mean Euclidean distance to the tau epicenter as well as random intercept. When extending this analysis to the cortex, we found a similar result pattern, with highest ^18^F-PI-2620 PET binding in those cortical regions that were most closely connected to subcortical tau epicenters (i.e. Q1) vs. lowest ^18^F- PI-2620 PET binding in those cortical regions that were most weakly connected to subcortical tau epicenters (i.e. Q4) in PSP-RS (Fig.5C, b/SE=0.036/0.008, p<0.001) and CBS (Fig.5E, b/SE=0.032/0.005, p<0.001). A topological frequency mapping of tau epicenters is shown in Fig.5H for PSP-RS and Fig.5I for CBS. Together, these findings support the view that tau spreading patterns follow the connectivity pattern of subcortical tau epicenters in PSP-RS and CBS.

**Figure 5:**
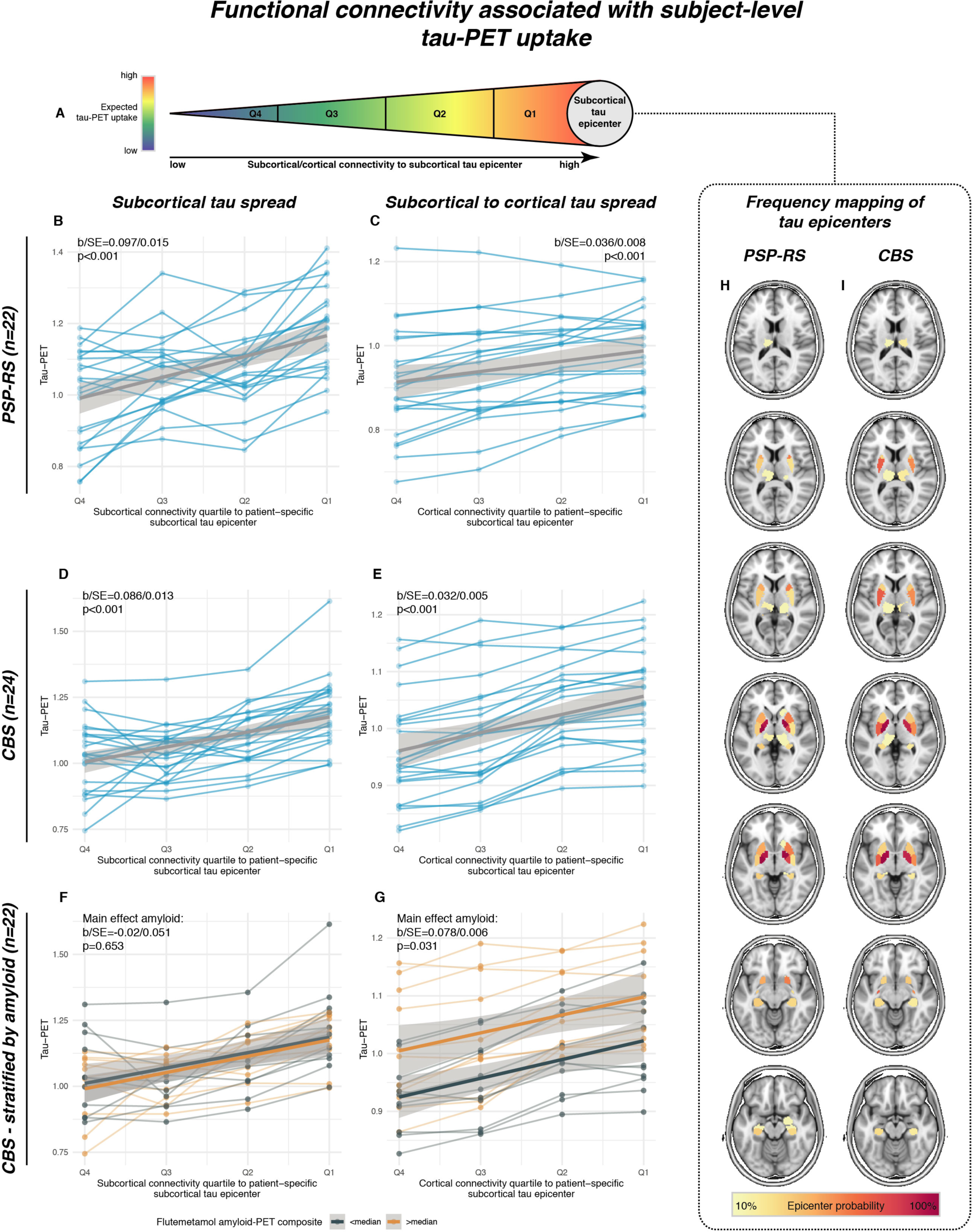
Using subject-level tau-PET data, we determined for each PSP-RS and CBS patient the subcortical tau epicenter (A), i.e. defined as 20% of ROIs with highest tau-PET SUVRs. The remaining ROIs were grouped for each subject into quartiles, depending on connectivity strength to the subject-specific tau epicenter. Highest tau- PET was expected for regions most closely connected to the tau epicenter (i.e. Q1) whereas lowest tau-PET was expected for ROIs only weakly connected to the tau epicenter. Subject-specific tau-PET data for subcortical Q1- Q4 ROIs (B&D) as well cortical Q1-Q4 ROIs (C&E) is shown, illustrating that tau-PET was highest in subcortical and cortical regions that are most closely connected to the subcortical tau epicenter (i.e. Q1), with gradual decreases across less strongly connected regions. For a subset of CBS patients (n=22), we further stratified these analyses by above vs. below median global amyloid-PET SUVRs (i.e. subthreshold amyloid, as all subjects were amyloid negative on visual read). Amyloid-stratified analyses illustrate that above median amyloid levels were not associated with elevated tau spread from subcortical epicenters to subcortical Q1-Q4 ROIs (F), but with increased tau spread from subcortical epicenters to cortical Q1-Q4 ROIs (G). All statistical indices (i.e. b-values, standard errors and p-values) were derived from linear mixed models, controlling for age, sex, study center, mean Euclidean distance of Q1-Q4 ROIs to the tau epicenter and random intercept. A probability mapping of subcortical epicenter locations is illustrated in panels H for PSP-RS and I for CBS patients.

### Subthreshold amyloid levels are associated with pronounced neocortical tau spread in CBS

In a subset of 22 CBS patients with available ^18^F-flutemetamol amyloid-PET (i.e. all rated Aβ- negative on expert visual read), we tested whether subthreshold Aβ levels are associated with enhanced subcortical to cortical tau spread. Here, we repeated the subject-level analyses illustrated in Figs.5D&E, this time including a main effect for high vs. low Aβ as defined via median split of global ^18^F-flutemetamol amyloid-PET SUVRs, intensity normalized to the pons. No difference in ^18^F-PI-2620 PET binding was found within subcortical tau epicenters between high and low Aβ groups (p>0.05). Further, the association between epicenter connectivity and ^18^F-PI-2620 PET binding in connected subcortical Q1-Q4 ROIs was the same across high/low Aβ groups (main effect of Aβ: b/SE=-0.02/0.051, p=0.653, Fig. 5F), controlling for age, sex, study site, Euclidean distance to the tau epicenter and random intercept. However, for cortical regions, we found that the high Aβ group showed overall higher ^18^F-PI-2620 PET binding across cortical Q1 to Q4 ROIs (main effect of Aβ: b/SE=0.078/0.006, p=0.031, Fig. 5G), yet with the same gradient of tau deposition from tau epicenters throughout connected regions, controlling for age, sex, study site, Euclidean distance to the tau epicenter and random intercept. This result pattern supports the view that subcortical to cortical tau spread is enhanced in the presence of subtle subthreshold Aβ accumulation in CBS.

### Functionally connected brain regions show correlated post-mortem tau levels in PSP

Lastly, we aimed to replicate the association between covariance in ^18^F-PI-2620 PET binding and functional connectivity using gold-standard post-mortem tau assessments. This analysis was motivated by missing autopsy validation of ^18^F-PI-2620 PET in 4R tauopathies. Thus, although binding patterns of ^18^F-PI-2620 closely reflect tau predilection sites of post-mortem studies, the in vivo data could still be driven by a parallel phenomenon not directly reflecting tau. The histopathological data of our study were derived from regional and cell-type-specific semi-quantitative AT8 (Munich sample) or PHF-1 (UPENN sample) stained tau assessments (i.e. for neuronal, astrocyte and oligodendrocyte tau) in two independent samples with confirmed PSP 4R tau pathology:^13^ Using the neuropathological probe extraction protocols, we reconstructed spatially matching bilateral ROIs from established cortical and subcortical anatomical atlases,^38, 40^ as shown in Fig.6A for the Munich-European consortium/collection sample (n=97, 16 ROIs) and Fig.6B for the UPENN sample (n=96, 9 ROIs). For these ROIs, we computed covariance in post-mortem stained neuronal tau levels, defined as the partial correlation between inter-regional neuronal tau, accounting for age at death and gender. Using the same ROIs shown in Figs.6A&B, we determined inter-regional functional connectivity in the sample of 69 cognitively normal, amyloid-PET and tau-PET negative ADNI subjects. As for the ^18^F-PI-2620 PET analyses, we tested the association between inter-regional functional connectivity and covariance in post-mortem tau, focusing particularly on neuronal tau which we hypothesized to be most strongly driven by connectivity-mediated tau spreading. As for ^18^F- PI-2620 PET, these analyses were controlled for inter-regional Euclidean distance, to ensure that associations between connectivity and tau covariance were independent of spatial proximity between ROIs. In line with the ^18^F-PI-2620 PET data, we found the expected association between functional connectivity and covariance in neuronal tau levels in the Munich-European consortium/collection (β=0.406, p<0.001, Fig.6G) and UPENN sample (β=0.565, p<0.001, Fig.6H). Again, these associations were confirmed by exact tests (p<0.001, beeswarm plots in Figs.6G&H), i.e. by comparing the actual β-value with a null-distribution of β-values obtained via repeating the analysis 1000 times using shuffled connectomes with preserved weight- and degree distribution. To test whether the association between functional connectivity and covariance in tau was strongest for neurons, we recomputed the above-described analyses with measures of neuronal, astroglial and oligodendroglial tau in 1000 bootstrapped samples that were randomly drawn from the overall samples. Plotting the resulting β-value distributions revealed that the associations between functional connectivity was indeed highest for neuronal tau, followed by oligodendrocyte and lastly astrocyte tau consistently across the Munich-European consortium/collection (Fig.6I) and UPENN sample (Fig.6J). Together, these post-mortem findings in two large independent samples replicate the in vivo ^18^F-PI-2620 PET findings, showing an association between functional connectivity and covariance in tau pathology, which is strongest for neuronal tau.

**Figure 6:**
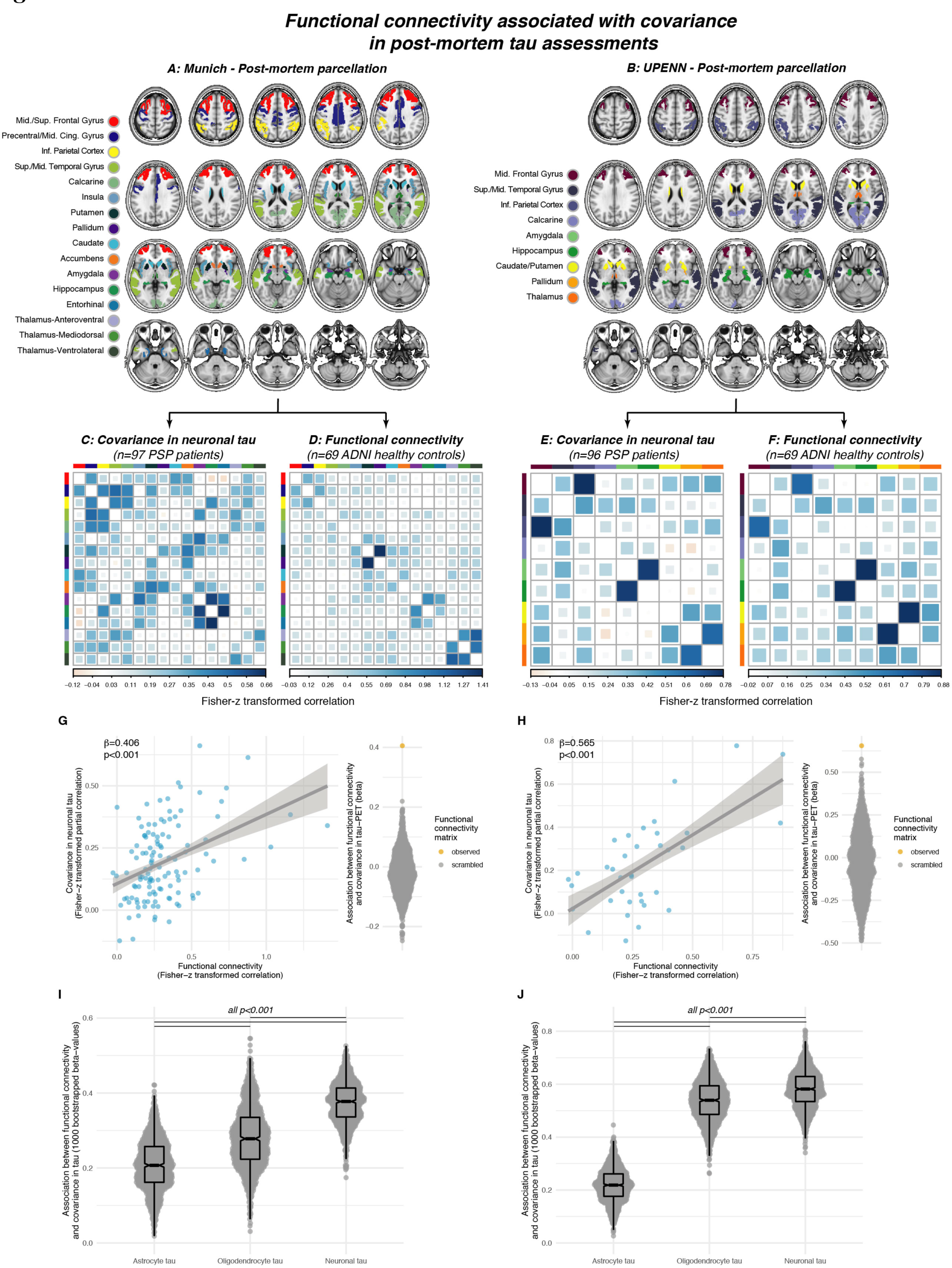
Association between functional connectivity and regional post-mortem tau assessments (i.e. AT8 staining) in two independent samples with histo-pathologically confirmed PSP. For each sample, covariance in neuronal tau levels was assessed among cortical and subcortical ROIs (A&B) using the methods illustrated in Fig. 3A, yielding a covariance in AT8-stained tau matrix of partial correlations accounting for age at death and sex. for the Munich (C) and UPENN sample (E). Using the same brain atlases (A&B), functional connectivity was determined based on resting-state fMRI data in the sample of n=69 healthy controls from ADNI (D&F). Scatterplots illustrate the association between functional connectivity and covariance in post mortem stained neuronal tau pathology for the Munich (G, AT8 tau staining) and UPENN (H, PHF-1 tau staining) sample. Standardized beta- and p-values were derived from linear regression controlling for Euclidean distance between ROIs. Robustness of the association in panels D&E was again tested by contrasting the beta-value derived from the association between the actual functional connectivity matrix with covariance in tau against beta-values derived from the same analyses repeated a 1000 times using scrambled functional connectivity matrices with preserved degree- and weight distribution (see beeswarm plots in panels G&H). The same analysis was repeated for cell-specific tau levels (i.e. astrocyte tau, oligodendrocyte tau, neuronal tau) using 1000 bootstrapping iterations, i.e. the association between functional connectivity and covariance in cell-specific tau was repeated on 1000 randomly drawn samples. The resulting beta-value distributions are shown for the Munich (I) and UPENN (J) sample, illustrating that the association between functional connectivity and covariance in tau is strongest for neuronal tau levels.

## DISCUSSION

Our major aim was to investigate whether functional connectivity is associated with the deposition patterns of tau pathology in 4R tauopathy patients. To this end, we combined template-based resting-state fMRI connectomics with i) in vivo tau-PET in 46 PSP-RS and CBS patients and ii) regional post-mortem tau assessments in two independent samples with histopathologically confirmed 4R tau PSP pathology (n=97/n=96). Using the next generation tracer ^18^F-PI-2620 for imaging tau pathology,^32^ we found elevated PET binding in clinically diagnosed CBS and PSP-RS patients, particularly in subcortical predilection sites of 4R tau pathology. Across CBS and PSP-RS, we show that functionally interconnected subcortical and cortical regions show correlated tau-PET levels. Moreover, we report that patient-level ^18^F-PI- 2620 PET patterns could be predicted by the seed-based connectivity patterns of subcortical tau epicenters, suggesting that gradual tau aggregation expands from subcortical tau starting sites throughout connected regions. In CBS, which is typically characterized by more widespread cortical tau,^28^ we show further that subthreshold Aβ-levels are associated with elevated cortical ^18^F-PI-2620 PET in regions that are closely connected to the subcortical tau epicenters. By translating the tau-PET vs. connectivity analysis approach to regionally sampled post-mortem data from two independent PSP patient samples, we replicated the association between in vivo-derived inter-regional connectivity and inter-regional covariance in AT8/PHF-1-stained tau pathology. Here, we could show further that the association between connectivity and covariance in post-mortem-assessed tau pathology was strongest for neuronal tau compared to glial tau levels, supporting the view that connectivity is particularly associated with trans- neuronal tau spread. Together, our findings provide comprehensive translational evidence for a key role of connectivity in the spreading of 4R tau pathology.^41, 42^ When assessing disease-associated tau-PET patterns, we found elevated ^18^F-PI-2620 PET binding in CBS/PSP-RS patients particularly in the basal ganglia, i.e. typical sites of 4R tau aggregation,^12, 13^ congruent with a previous ^18^F-PI-2620 PET study in a partly overlapping sample.^32^ Further, the epicenters of ^18^F-PI-2620 PET spatially matched the earliest signs of tau pathology in the pallido-nigrolusyian axis as detected in post-mortem analyses of 4R tauopathy patients with various clinical phenotypes.^13^ CBS patients, which are characterized by a clinically mixed subcortical/cortical phenotype showed more widespread cortical ^18^F-PI-2620 PET elevations than PSP-RS, in line with post-mortem observations.^28^ Together, the ^18^F-PI- 2620 PET patterns observed in PSP-RS and CBS are congruent with histopathologically observed 4R tau patterns in CBD/PSP.^12, 13^

Nevertheless, we note as a limitation that in vivo and post-mortem data were not assessed in the same patients in the current study. Thus, although there is growing evidence for ^18^F-PI-2620 binding to 4R tau,^32, 43^ the observed signal could still derive from a source closely paralleling 4R tau. Regarding our main finding, we observed that functionally connected brain regions exhibit correlated ^18^F-PI-2620 PET levels in CBS and PSP-RS. This result echoes previous evidence of covarying cross-sectional tau-PET levels and longitudinal tau-PET accumulation rates among functionally interconnected regions in AD, suggesting a consistent association between tau spreading and brain connectivity across different tauopathies.^44–46^ This interpretation is also supported by preclinical work, showing that pathological tau species from 4 repeat (i.e. CBD/PSP) and mixed 3/4 repeat tauopathies (i.e. AD) consistently trigger trans- cellular tau spread across connected brain regions while recapitulating disease-specific neuropathological hallmark features (e.g. neuronal tau in AD vs. neuronal and glial tau in CBD/PSP).^20, 22, 47^ Importantly, the association between connectivity and covariance in ^18^F-PI- 2620 PET binding was detected for subcortical 4R tau predilection sites but also for cortico-cortical and subcortico-cortical connections. This indicates that associations between connectivity and tau spreading are not spatially restricted to particularly vulnerable brain regions, in line with previous evidence in AD patients^39, 44, 45^ and preclinical tauopathy models.^20, 22, 47^

In a critical validation step, we were able to translate these in vivo tau-PET vs. connectivity analyses to post-mortem tau assessments in two pre-existing datasets of patients with histopathologically confirmed PSP.^13^ In these unique samples with gold-standard regional tau assessments, we found a highly consistent result pattern with strong associations between in vivo-assessed inter-regional functional connectivity and inter-regional correlations of AT8 or PHF-1 stained tau levels. Importantly, these associations were replicated across both independent post-mortem samples with regionally different post-mortem sampling (see Figs.6A&B), supporting the robustness of our findings. Even more important, cell-type-specific tau assessments confirmed the hypothesis that the association between connectivity and covariance in tau was strongest for neuronal tau pathology when compared to astroglial and oligodendrocyte-specific tau, thus supporting neuronal connectivity as the main driver of trans-synaptic tau propagation.^27^ Consistent across both post-mortem samples, the second strongest association between connectivity and tau covariance was observed for oligodendrocytes, i.e. the main constituents of myelin sheaths along axonal tracts. Previous work in mice has shown that 4R oligodendrocyte tau propagates along white matter tracts, even in the absence of neuronal tau pathology, suggesting oligodendrocyte tau propagation along anatomically interconnected pathways.^48^ A recent study evaluating cell-specific sequences showed more overlap of the patterns of neuronal and oligodendrocytic tau deposition, and albeit early steps of astrocytic tau deposition deviates from the neuronal, it converges in later steps.^13^ Astrocytic tau accumulation may be seen without local neuronal tau pathology in regions with high connectivity to regions with existing neuronal tau pathology,^49^ which together with a preclinical study that observed astrocytic tau pathology only in the presence of neuronal tau,^48^ are in line with the observation that astrocytes phagocytose neuronally released tau^20^, either from local neurons or projecting from other regions.^50^ Altogether, these may explain why connectivity and astrocytic tau were associated, albeit to a weaker extent compared to neuronal and oligodendrocytic tau. Thus, these preclinical findings converge with our result pattern of strongest associations between connectivity and neuronal tau, followed by oligodendrocyte and astrocyte tau. In summary, our findings provide compelling post-mortem replication of an association between connectivity and 4R tau spreading.

Importantly, all associations between connectivity and covariance in post-mortem tau or in vivo ^18^F-PI-2620 PET were detected while statistically controlling for inter-regional Euclidian distance, highlighting that the association between tau and connectivity is not driven by mere spatial proximity. This result pattern supports the view that tau spread is driven by connectivity and not proximity,^26, 51, 52^ in line with pre-clinical studies, showing that cerebral tau injections trigger tau spread to connected rather than spatially adjacent regions.^20, 47^

In the ^18^F-PI-2620 PET sample, we further show that brain regions with highest tau (i.e. tau epicenters) are closely functionally connected to other high tau regions, whereas low tau regions are most closely connected to other low tau regions. These findings are congruent with post-mortem tau staging schemes, showing that 4R tau pathology is initially confined to circumscribed epicenters from where it progresses gradually throughout the brain.^13^ Further supporting this, we found that connectivity patterns of subject-level tau-PET epicenters predicted a gradient of tau deposition, with highest tau in subcortical/cortical regions that were most closely connected to the tau epicenter, while tau levels were gradually lower in farther connectivity-based distance to the tau epicenter. In line with findings in other tauopathies such as AD,^39, 44, 46, 53^ these results suggest tau spreading patterns in 4R tauopathies are determined by the connectivity pattern of the tau epicenter, thereby strongly supporting the concept of connectivity-based tau spreading. Importantly tau spreading concepts applied to PET imaging could facilitate patient-tailored precision medicine since they facilitate prediction of the future accumulation pattern of tau pathology and the progression/development of symptoms on a patient-level.^39^ By implication, identifying sites that are vulnerable to tau spreading could also serve as a imaging-guided primary read-out in tau targeting trials of 4R tauopathies.

In an exploratory sub-analysis in CBS patients with available global amyloid-PET data, we found further that subthreshold Aβ-levels were associated with higher cortical tau-PET-levels following the same connectivity gradient of subcortical tau epicenters (Fig.5G). A potential explanation is that subtly elevated Aβ levels (i.e. all CBS patients were by definition Aβ- negative), which have been previously shown to accelerate AD-typical tau spreading,^54^ also accelerate subcortical to cortical tau spreading in CBS patients. As a limitation, we note that different binding characteristics of ^18^F-PI-2620 to 3/4R and 4R tau isoforms could also explain our observation since the Aβ status impacts the predominant tau isoform and tau isoform shifts.^55^ Yet, this finding is preliminary and requires further systematical investigation by larger dedicated studies.

Several limitations should be considered when interpreting our results. First, the ^18^F-PI-2620 PET cohort includes PSP-RS and CBS patients with clinically diagnosed 4R tauopathies, since biomarker-based 4R tauopathy diagnosis is not yet clinically established.^17, 33^ Importantly, ^18^F- PI-2620 PET patterns in PSP-RS and CBS groups matched the tau patterns that are typically observed in histopathological assessments.^12, 13^ Further, subcortical ^18^F-PI-2620 PET was significantly higher relative to controls, thus supporting the view that the ^18^F-PI-2620 PET signal in these 4R tau predilection sites is not systematically confounded by off-target binding.^32^ In addition, all patients with CBS were rated Aβ negative as determined via cerebrospinal fluid or amyloid-PET assessments, suggesting that elevated ^18^F-PI-2620 PET is not primarily explained by AD-typical 3R/4R tau.^56, 57^ Second, the current study focused on functional connectivity, i.e. shared inter-regional BOLD activity, which is to a large degree but not entirely matched by structural connectivity as assessed via diffusion MRI.^58^ This structural/functional connectivity mismatch is partly determined by technical limitations of MRI-based tractography to detect crossing-fibers or short-range cortico-cortical connections.^59^ On the other hand, the slow temporal resolution of fMRI may introduce connectivity between regions without direct but rather indirect multi-synaptic connections.^60^ Thus, our results on tau covariance vs. connectivity likely reflect a mixture of direct and indirect connections. However, the advantage of functional MRI in the current study is the possibility to map connections between spatially adjacent subcortical nuclei, where connections are not accessible with diffusion-based tractography. In addition, the current study used connectivity templates derived from healthy individuals. Thus, it remains to be determined by future studies whether subject-specific connectivity differences (i.e. connectome fingerprinting) contribute to heterogeneity in tau spreading patterns. Third, the current study is cross-sectional in nature and thus does not assess the association between connectivity and the spatio-temporal progression of tau pathology. In order to test the predictive value of connectivity for regional changes in tau-PET, longitudinal tau-PET studies are necessary, which will be conducted as soon as large enough data are available.

In conclusion, the current study demonstrates a close link between 4R tau deposition patterns and connectivity, thereby supporting the concept of trans-neuronal tau spreading in 4R tauopathies.^18, 27, 42^ A clear strength of the current study is the translational study design with independent validation of the association between inter-regional functional connectivity and covariance in tau pathology across in vivo ^18^F-PI-2620 PET and cell-specific post-mortem histopathological tau assessments in multiple independent samples. Since tau pathology is closely associated with disease severity and clinical symptom expression,^5, 12, 13^ our results suggest that interventions that target tau spreading are potentially key therapeutic strategies.

## METHODS

### Tau-PET Sample

We included 61 subjects recruited at four sites (Munich & Leipzig, Germany; Melbourne, Australia; New Haven, United States), including 15 cognitively normal individuals (i.e. without evidence of cognitive decline, any motor symptoms or cerebral tau pathology), 24 patients with clinical diagnosis of possible or probable cortico-basal syndrome (CBS) and 22 patients with clinical diagnosis of PSP-RS. CBS diagnosis was made according to the revised Armstrong Criteria of probable CBS^17^ or the Movement Disorders Society criteria of possible PSP with predominant CBS.^33^ PSP-RS was diagnosed following state-of-the-art diagnostic criteria.^33^ All patient data derive from a PSP cohort recruited in Munich and Leipzig^32^ and a CBS cohort recruited in Munich.^36^ Inclusion criteria for the current study were age above 45 years, stable pharmacotherapy for at least one week before the tau-PET examination, negative family history for Parkinson’s and Alzheimer’s disease and availability of 3D T1-weighted structural MRI. Exclusion criteria were severe neurological or psychiatric disorders other than PSP and CBS or positive AI status, as determined via expert visual read of 18-F-Flutemetamol or 18-F-florbetaben amyloid-PET or by cerebrospinal fluid analyses of Aβ levels using locally established cut-offs (i.e. Aβ42/40-ratio < 5.5% or Aβ1-42<375pg/ml). The 18-F-PI2620 PET imaging protocol was approved by local ethics committee of the LMU Munich. Written informed consent was obtained from all participants. The full study protocol including all samples and all PET data analyses were approved by the local ethics committee (LMU Munich, application numbers 17-569 and 19-022) and the German radiation protection (BfS-application: Z 5 - 22464/2017-047-K-G) authorities. The study was carried out according to the principles of the Helsinki Declaration. The post-mortem examinations of brain tissue in PSP patients were approved by the ethics committee of the medical faculty of the university of Marburg, Germany. All work complied with ethical regulations for work with human participants.

### Neuroimaging acquisition

All structural MRI data was collected on 3T scanners using 3D MPRAGE and MP2RAGE sequences. Radiosynthesis of 18-F-PI-2620 was achieved by nucleophilic substitution on a butyloxycarbonyl-protected nitro precursor using an automated synthesis module (Synthera, IBA Louvain-la-neuve, Belgium). The protecting group was cleaved under radiolabeling conditions. The product was purified by semipreparative high performance liquid chromatography, resulting in radiochemical purity of >97%. Non-decay corrected yields were about 30% with a molar activity of 3·10^6^ GBq/mmol at the end of synthesis. 18-F-PI-2620 PET imaging in combination with computed tomography (CT) or magnetic resonance (MR) was performed in a full dynamic setting (minimum scan duration: 0–60 min post-injection) using pre-established standard PET scanning parameters at each site: In Munich, Germany, dynamic tau-PET was acquired on a Siemens Biograph True point 64 PET/CT (Siemens, Erlangen, Germany) or a Siemens mCT scanner (Siemens, Erlangen, Germany) in 3D list-mode over 60min and reconstructed into a 336x336x109 matrix (voxel size: 1.02×1.02×2.03mm^3^) using the built-in ordered subset expectation maximization (OSEM) algorithm with 4 iterations, 21 subsets and a 5mm Gaussian filter. A low dose CT was used for attenuation correction. In Leipzig, Germany, dynamic tau-PET was acquired on a hybrid PET/MR system (Biograph mMR, Siemens Healthineers, Erlangen, Germany) in 3D list-mode over 60 min and reconstructed into a 256x256 matrix (voxel size: 1.00×1.00×2.03mm^3^) using the built-in ordered subset expectation maximization algorithm with 8 iterations, 21 subsets and a 3mm Gaussian filter. For attenuation correction, the vendor-provided HiRes method was employed, combining the individual Dixon attenuation correction approach with a bone attenuation template.^61^ For tau-PET data of control subjects imaged in New Haven, US, dynamic PET was acquired on a Siemens ECAT EXACT HR+ camera from 0-90 and 120-180 min. Images were reconstructed in a 128 x 128 matrix (zoom=2, pixel size of 2.574 mm x 2.574 mm) with an iterative reconstruction algorithm (OSEM 4 iterations, 16 subsets) and a post-hoc 5mm Gaussian filter. Standard corrections for random, scatter, system dead time and attenuation provided by the camera manufacturer were performed. 18-F-PI-2620 PET assessments of control subjects in Melbourne was performed on a Philips Gemini TF 64 PET/CT (Philips, Eindhoven, The Netherlands). PET images were acquired dynamically from 0-60 min and 80- 120 min post-injection. Images were reconstructed using LOR-RAMLA and CT attenuation correction was performed. Images were binned into a 128x128x89 matrix (voxel size: 2.00×2.00×2.00 mm^3^).

The injected dose was 168 to 334 MBq, applied as a bolus injection. Site-specific attenuation correction ensured multi-site harmonization of data acquisition.^61^ Further, data from Hofmann brain phantoms were used to obtain scanner-specific filter functions which were consequently used to generate images with a similar spatial resolution for voxel-wise analyses (full-width-at-half-maximum=9x9x10mm; determined by the scanner in New Haven), following the Alzheimer’s Disease Neuroimaging Initiative image harmonization procedure.^62^ Resulting smoothing factors were 3.5x3.5x7.0mm for Munich, 6.0x6.0x6.0mm for Leipzig, and 4.0x4.0x4.0mm for Melbourne. All dynamic datasets were visually checked for artifacts and motion-corrected using rigid-registration. Mean SUV images were obtained for 20-40 min time frames to obtain SUVR images which show comparable performance in signal sensitivity and were less subject to artifacts when compared to 0-60 min or 0-40 min DVR images.^63^

### Structural MRI and tau-PET preprocessing

All structural MRI and PET data was processed using the Advanced Normalization Tools (ANTs) toolbox (http://stnava.github.io/ANTs/). In an initial step, 18-F-PI-2620 PET images were rigidly co-registered to native-space T1-weighted MRI images. For T1-weighted structural MRI data we performed bias field correction, brain extraction, and segmentation into grey-matter, white matter and cerebrospinal fluid tissue maps using the ANTs cortical thickness pipeline. Brain extracted T1-weighted images were non-linearly normalized to MNI space

(2mm isotropic voxels) using ANTs high-dimensional warping algorithm.^64^ By combining the rigid 18-F-PI-2620 PET to T1-native space and non-linear T1 to MNI space spatial normalization parameters, all brain-atlas derived ROI data was transformed from MNI space back to PET native space, including the Tian 32 ROI subcortical brain atlas^38^ (Fig.1H), the Schaefer 200 ROI cortical brain atlas^37^ (Fig.1G), as well as the inferior cerebellar reference ROI for intensity normalization of tau-PET.^65^ All brain atlas data and the inferior cerebellar reference ROI were further masked with binary subject-specific grey matter maps in order to restrict later extraction of ROI-mean values to grey matter regions.

Tau-PET images were intensity normalized to the mean tracer uptake of the inferior cerebellar grey, to determine standardized uptake value ratio (SUVR). Mean tau-PET SUVR values were extracted for each subject for the 32 subcortical and 200 cortical ROIs. For voxel-wise analyses, subject-specific tau-PET SUVR images were warped to MNI space by combining the linear PET to T1 and non-linear T1 to MNI transformation parameters, followed by spatial smoothing with site-specific smoothing kernels. Usage of an alternative reference ROI (i.e. eroded white matter) yielded congruent analyses with those presented in the manuscript.

### Post-mortem sample

To replicate tau-PET vs. connectivity associations using post-mortem assessments of tau pathology, we included histopathological tau data from two independent samples, including n=97 PSP subjects recruited, sampled and examined at several different sites across Europe with centralized final analysis at the Department of Neuropathology, LMU, in Munich and n=96 PSP subjects from the University of Pennsylvania. An in-depth description of data selection and data acquisition has been published previously.^13^ Cases were selected based on presence of neurofibrillary tangles in the subthalamic nucleus, substantia nigra and pallidum, as well as based on the presence of tufted astrocytes in the striatum and frontal cortex.^3, 66^

Extraction of neuropathological samples followed a standardized procedure as described previously.^13^ Formalin fixed and paraffin-embedded tissue blocks from the PSP cases were evaluated using tau immunostaining with the anti-tau AT8 antibody (Ser202/Thr205, 1:200, Invitrogen/Thermofischer, MN1020, Carlsbad, USA) for the Munich sample, and with anti-tau PHF-1 (Ser396/Ser404, 1:2000) for the UPENN sample. For each sample, we included data from post-mortem sampled regions of interest, which were judged accessible with functional MRI, resulting in 16 ROIs for the Munich sample and 9 ROIs for the UPENN sample. Neuropathological ROIs were reconstructed in MNI space based on the neuropathological examination protocol using pre-defined ROIs from established anatomical brain atlases (Figs.6A&B).^38, 40^ Neuronal tangle pathology was graded for each region in a semiquantitative score (none=0, mild=1, moderate=2, severe=3).

### Assessment of covariance in tau-PET and post-mortem tau

We assessed the inter-regional covariance in 18-F-PI-2620 PET SUVRs (see Fig.2A for an analysis flow-chart) for 22 PSP-RS and 24 CBS patients. The analysis pipeline was adopted from previous studies using this approach to determine FDG-PET covariance (i.e. metabolic covariance), grey matter covariance (i.e. structural connectivity) or 18-F-AV1451 PET covariance across brain regions.^44, 45, 67, 68^ First, we computed the mean tau-PET uptake within each of the 200 cortical (Fig.1G) and 32 subcortical (Fig.1H) ROIs for each subject (Fig.2, A(i)). Next, we vectorized mean ROI SUVR values to subject-specific 232-element vectors (Fig.2, A(ii)). Using these 232-element 18-F-PI-2620 PET SUVR vectors, we then assessed across subjects the pairwise ROI-to-ROI partial correlation of 18-F-PI-2620 PET uptake (Fig.2, A(iii)), adjusting for subject-specific age, gender and PET imaging and MRI protocol as potential confounds. This analysis resulted in a 232x232 sized 18-F-PI-2620 PET covariance matrix each for each group (i.e. PSP, CBS). Within this 18-F-PI-2620 PET covariance matrix, autocorrelations were set to zero and all correlations were Fisher-z transformed.

An equivalent approach was used for post-mortem tau data. For each sample, we determined the inter-regional partial correlation of post-mortem AT8-stained tau pathology across the 16 ROIs of the Munich-European consortium/collection sample (Fig.6A) or 9 ROIs of the UPENN sample (Fig.6B), adjusting for age at death and gender, yielding a covariance in post-mortem tau matrix (Figs.6C&E). Again, autocorrelations within this matrix were set to zero and all remaining correlations were Fisher-z transformed.

### Functional connectivity assessment

For assessing functional connectivity, we used resting-state fMRI data from 69 cognitively normal controls of the ADNI cohort. These subjects were selected based on absence of objective or subjective signs of cognitive impairment and had no evidence of clinically relevant cerebral amyloid or tau pathology, as indicated by negative ^18^F-florbetapir amyloid-PET (i.e. global SUVR<1.11) ^69^ and negative ^18^F-flortaucipir tau-PET scans (i.e. global SUVR<1.3) ^34^.

MRI scans were obtained on Siemens scanners using unified scanning protocols. T1-weighted structural MRI was recorded using a MPRAGE sequence with 1mm isotropic voxel-space and a TR=2300ms. For functional MRI, for each subject a total of 200 resting-state fMRI volumes were recorded using a 3D echo-planar imaging (EPI) sequence in 3.4mm isotropic voxel resolution with a TR/TE/flip angle=3000/30/90°.

All images were inspected for artifacts prior to preprocessing. Using ANTs, T1-weighted structural MRI images were bias corrected, brain extracted, segmented and non-linearly spatially normalized to MNI space.^64^ Functional EPI images were slice-time and motion corrected (i.e. realignment to the first volume) and co-registered to the native T1 images. Using rigid-transformation parameters, T1-derived grey-matter, white matter and cerebrospinal fluid segments were transformed to EPI space. To denoise the EPI images, we regressed out nuisance covariates (i.e. average white matter and cerebrospinal fluid signal and motion parameters estimated during motion correction), removed the linear trend and applied band-pass filtering with a 0.01-0.08Hz frequency band in EPI native space. To further minimize the impact of motion which may compromise FC assessment,^70^ we performed motion scrubbing, where we censored volumes that showed a frame-wise displacement of >1mm, as well as one prior and two subsequent volumes. In line with our previous work,^71^ only subjects for whom less than 30% of volumes had to be censored were included in the current study. Note that we did not spatially smooth the functional images during preprocessing to avoid signal spill-over between adjacent brain regions that may artificially enhance functional connectivity between adjacent brain regions during ROI based connectivity assessment.

To determine functional connectivity, we warped the 232 cortical and subcortical ROIs for tau-PET analyses (Fig. 1G&H) as well as the post-mortem ROIs for the Munich-European consortium/collection and UPENN sample (Figs.6A&B) to the denoised and preprocessed fMRI images in native EPI space, by combining the linear EPI to T1 and non-linear T1 to MNI transformation parameters. ROI maps in EPI space were masked with subject-specific grey matter. Fisher-z transformed Pearson-Moment correlations between time-series averaged across voxels within an ROI were determined to assess subject-specific functional connectivity matrices. Functional connectivity data was averaged across all 69 ADNI subjects in order to determine group-average functional connectivity matrices for the in-vivo tau-PET analyses as well as for the post-mortem analyses.

For the group-average functional connectivity matrices, we further determined 1000 null-models of functional connectivity respectively, by shuffling the functional connectivity matrices while preserving the overall degree- and weight-distribution, using the null_model_und_sign.m function of the brain connectivity toolbox (https://sites.google.com/site/bctnet/).

### Statistics

Sample demographics were compared between the groups using ANOVAs for continuous measures and Chi-squared tests for categorical measures. Voxel-wise comparisons in 18-F-PI- 2620 PET SUVRs were determined on spatially-normalized and smoothed tau-PET images using ANCOVAs in SPM12, controlling for age, gender and study site, applying a voxel-wise alpha threshold of 0.005 and a cluster extent threshold of >100 spatially contiguous voxels.

To test the association between functional connectivity and covariance in 18-F-PI-2620 PET, we used linear regression with covariance in 18-F-PI-2620 PET as a dependent variable and ADNI-derived group-average functional connectivity as an independent variable, controlling for inter-regional Euclidean distance (i.e. distance between ROI-specific centers of mass). This analysis was stratified by CBS and PSP-RS and conducted for subcortical ROIs only (i.e. primary analysis) as well as for the whole brain (i.e. secondary analysis). The same analysis was performed for post-mortem tau assessments in the Munich-European consortium/collection and UPENN dataset, using regional semi-quantitative tau data (i.e. 16 ROIs in the Munich sample vs. 9 ROIs in the UPENN sample) rather than tau-PET. To determine the robustness of the association between connectivity and covariance in 18-F-PI-2620 PET/post-mortem tau, the analysis was repeated 1000 times using shuffled connectomes with preserved weight- and degree distribution to obtain a distribution of “null-model” β-values. We then performed an exact test, i.e. we determined the probability of null-distribution derived β-values surpassing the true β-value. For post-mortem data, we further assessed cell-type-specific associations between tau covariance (i.e. neuronal, astroglial and oligodendroglial tau covariance) and functional connectivity. To this end we computed regression-based association between covariance in tau and functional connectivity for each cell type, based on 1000 bootstrapped samples that were randomly drawn from the respective sample (i.e. UPENN or Munich). The resulting β-value distributions reflecting the association between connectivity and cell-type-specific tau covariance were compared between cell-types using paired t-tests.

Using 18-F-PI-2620 PET, we further tested, whether functional connectivity patterns of tau epicenters (i.e ROIs with highest tau) are predictive of 18-F-PI-2620 PET binding in remaining brain regions. Again, this analysis was determined stratified by group (i.e. PSP-RS vs. CBS) and conducted for subcortical ROIs only (i.e. primary analysis) as well as for the whole brain parcellation (i.e. secondary analysis). Specifically, we determined group-level 18-F-PI-2620 PET binding, and tested whether higher seed-based connectivity of the epicenter ROI was associated with higher 18-F-PI-2620 PET binding in the remaining ROIs, using linear regression controlling for between-ROI Euclidean distance. The same analysis was performed for the ROI with lowest 18-F-PI-2620 PET binding (i.e. coldspot), for which we assumed that higher connectivity is associated with lower 18-F-PI-2620 PET binding in the remaining regions. In an iterative next step, we repeated this analysis across all ROIs and determined the association between the 18-F-PI-2620 PET level in the seed ROI against the regression-derived

β-value of the association between seed-based connectivity and 18-F-PI-2620 PET binding in remaining ROIs. As in our previous studies,^44, 45^ we expected that ROIs with higher tau-PET binding should be connected to other ROIs with a high binding level (i.e. as reflected in a positive β-value), whereas regions with low tau-PET binding should be connected to other ROIs with a low binding level (i.e. as reflected in a negative β-value).

For subject-level analyses, we adopted our pre-established approach^39^ and determined subject-level epicenters as 20% of ROIs with highest tau-PET binding (see Fig.5H&I for tau epicenter probability maps in PSP-RS and CBS). The remaining non-epicenter ROIs were grouped for each subject into 4 quartiles, based on the connectivity strength to the epicenter (i.e. ROIs grouped in Q1 show highest connected to the tau epicenter, vs. ROIs grouped in Q4 show weakest connectivity to tau epicenters). Note that Q1-Q4 ROIs were determined separately for subcortical and cortical regions. Mean 18-F-PI-2620 PET binding was assessed for each subjects’ subcortical/cortical Q1-Q4 ROIs. Using linear mixed models, we then tested the association between connectivity strength (i.e. Q1-Q4) and subcortical or cortical 18-F-PI-2620

PET binding in Q1-Q4 ROIs, controlling for age, sex, study site, mean Euclidean distance to the epicenter as well as random intercept. In an exploratory analysis in the CBS group, we repeated this subject-level analysis while adding an additional main effect for global ^18^F- Flutemetamol amyloid-PET levels stratified at the median, to determine whether subthreshold levels of Aβ were associated with increased subcortical to cortical tau spread. Altering the definition of epicenters (i.e. 10%, 15%, 20%, 25% or 30% of ROIs with highest 18-F-PI-2620 PET binding) did not change the result pattern. All analyses were computed using R statistical software (r-project.org).

### Data availability statement

The data that support the findings of this study are available on reasonable request from the corresponding author.

## Acknowledgements

We wish to thank the patients and their families, without whose support and altruism this research would not have been possible. Further, we would like to acknowledge Prof. Christian Haass (DZNE, Munich) for his consultation on study design, the European Reference Network for Rare Neurological Diseases - Project ID No 739510 and the Neurological Tissue Bank of the Biobanc-Hospital Clinic-IDIBAPS for sample and data procurement. This work was supported by the NIH (P30AG010124, P01AG066597, U19AG062418, R01-NS109260), the National Center for Advancing Translational Sciences (TL1TR001880), the German Research Foundation (DFG; SCHR 774/5-1, DFG, INST 409/193-1 FUGG) the SyNergy excellence cluster (EXC 2145/ID 390857198). NF was supported by the Hertie foundation for clinical neurosciences. GGK is supported by the Rossy Foundation and Safra Foundation. GH was funded by the Deutsche Forschungsgemeinschaft (DFG, HO2402/18-1 MSAomics), the German Federal Ministry of Education and Research (BMBF, 01EK1605A HitTau), VolkswagenStiftung and Lower Saxony Ministry for Science (Niedersächsisches Vorab), Petermax-Müller Foundation (Etiology and Therapy of Synucleinopathies and Tauopathies). MB was supported by the Hirnliga e.V. (Manfred-Strohscheer-Stiftung) and by the Alzheimer Forschung Initiative e.V. (grant number #19063p).

